# A randomized, double-blind, controlled trial of convalescent plasma in adults with severe COVID-19

**DOI:** 10.1101/2021.03.12.21253373

**Authors:** Max R. O’Donnell, Beatriz Grinsztejn, Matthew J. Cummings, Jessica Justman, Matthew R. Lamb, Christina M. Eckhardt, Neena M. Philip, Ying Kuen Cheung, Vinay Gupta, Esau João, Jose Henrique Pilotto, Maria Pia Diniz, Sandra Wagner Cardoso, Darryl Abrams, Kartik Rajagopalan, Sarah Borden, Allison Wolf, Leon Claude Sidi, Alexandre Vizzoni, Valdilea G. Veloso, Zachary C. Bitan, Dawn E. Scotto, Benjamin J. Meyer, Samuel D. Jacobson, Alex Kantor, Nischay Mishra, Lokendra V. Chauhan, Elizabeth Stone, Flavia Dei Zotti, Francesca La Carpia, Krystalyn E. Hudson, Stephen A. Ferrera, Joseph Schwartz, Brie Stotler, Wen-Hsuan Lin, Sandeep Wontakal, Beth Shaz, Thomas Briese, Eldad A. Hod, Steven L. Spitalnik, Andrew Eisenberger, W. Ian Lipkin

## Abstract

**Background:** Although convalescent plasma has been widely used to treat severe coronavirus disease 2019 (COVID-19), data from randomized controlled trials that support its efficacy are limited.

**Objective:** To evaluate the clinical efficacy and safety of convalescent plasma among adults hospitalized with severe and critical COVID-19.

**Design:** Randomized, double-blind, controlled, multicenter, phase 2 trial conducted from April 21^st^ to November 27^th^, 2020.

**Setting:** Five hospitals in New York City (NY, USA) and Rio de Janeiro (Brazil).

**Participants:** Hospitalized patients aged ≥18 years with laboratory-confirmed COVID-19, infiltrates on chest imaging and oxygen saturation ≤ 94% on room air or requirement for supplemental oxygen, invasive mechanical ventilation, or extracorporeal membrane oxygenation.

**Intervention:** Participants were randomized 2:1 to a single transfusion of either 1 unit of convalescent or normal control plasma.

**Measurements:** The primary outcome was clinical status at 28 days, measured using an ordinal scale and analyzed using a proportional odds model in the intention-to-treat population.

**Results:** Of 223 participants enrolled, 150 were randomized to receive convalescent plasma and 73 to normal control plasma. At 28 days, no significant improvement in clinical status was observed in participants randomized to convalescent plasma (with an odds ratio (OR) of a 1-point improvement in the scale: 1.50, 95% confidence interval (CI) 0.83-2.68, p=0.180).

However, 28-day mortality was significantly lower in participants randomized to convalescent plasma versus control plasma (19/150 [12.6%] versus 18/73 [24.6%], OR 0.44, 95% CI 0.22-0.91, p=0.034). The median titer of anti-SARS-CoV-2 neutralizing antibody in infused convalescent plasma units was 1:160 (IQR 1:80-1:320). In a subset of nasopharyngeal swab samples (n=40) from Brazil that underwent genomic sequencing, no evidence of neutralization-escape mutants was detected. Serious adverse events occurred in 39/147 (27%) participants who received convalescent plasma and 26/72 (36%) participants who received control plasma.

**Limitations:** Some participants did not receive high-titer convalescent plasma.

**Conclusion:** In adults hospitalized with severe COVID-19, use of convalescent plasma was not associated with significant improvement in 28 days clinical status. The significant reduction in mortality associated with convalescent plasma, however, may warrant further evaluation.

**Registration:** ClinicalTrials.gov, NCT04359810

**Funding:** Amazon Foundation

**Clinical Trial Registration:** ClinicalTrials.gov Identifier: NCT04359810

## Introduction

As of March 4^th^, 2021, over 115 million cases of coronavirus disease 2019 (COVID-19) associated with severe acute respiratory syndrome coronavirus-2 (SARS-CoV-2) had been reported worldwide (1). Available data suggest that approximately 10-25% of patients with SARS-CoV-2 infection develop severe COVID-19 characterized primarily by pneumonia and in a subset, acute respiratory distress syndrome (ARDS) (2–4) and among severe cases, mortality occurs in 39-49% (2,4).

Following the emergence of SARS-CoV-2, convalescent plasma was proposed as a rapidly scalable therapeutic to prevent or mitigate severe illness through virus neutralization or antibody-dependent immunomodulation (5). During recent epidemics of emerging respiratory viruses such as SARS-CoV, H5N1 and 2009 H1N1 influenza, observational and non-randomized studies reported improved clinical outcomes and minimal adverse effects associated with use of convalescent plasma in severely ill patients (6). In patients with severe COVID-19, observational studies have suggested possible clinical efficacy and safety using convalescent plasma, primarily among patients not receiving invasive mechanical ventilation (IMV) and those with shorter durations of illness (7–10). Despite these signals, data from randomized controlled trials supporting use of convalescent plasma in hospitalized patients with COVID-19 are limited. Open-label trials from India and China reported no significant improvements in clinical outcomes among patients hospitalized with severe COVID-19; the latter was substantially underpowered (11,12). A recent double-blind, placebo-controlled trial in Argentina reported no improvement in clinical outcomes with use of convalescent plasma among adults hospitalized with severe COVID-19, including among subgroups stratified by illness duration and clinical severity (13).

In the United States and Brazil, approximately 28.7 and 10.7 million cases of Covid-19 have been reported as of March 4^th^, 2021, respectively (1). Given the lack of effective medical therapies against SARS-CoV-2, we conducted a randomized, double-blind, controlled phase 2 clinical trial to evaluate the clinical efficacy and safety of convalescent plasma among adults hospitalized with severe and critical COVID-19 in New York City and Rio de Janeiro.

## Methods

### Study Design

This was an investigator-initiated, randomized, double-blind, controlled trial to evaluate the efficacy and safety of convalescent plasma among adults hospitalized with severe COVID-19. The trial was conducted at five sites in New York City (USA) and Rio de Janeiro (Brazil) and was coordinated by Columbia University. Study sites included two hospitals affiliated with New York-Presbyterian Hospital/Columbia University Irving Medical Center (CUIMC) in northern Manhattan (Milstein and Allen Hospitals) and three sites in Rio de Janeiro (Instituto Nacional de Infectologia Evandro Chagas, Hospital Federal dos Servidores do Estado, and Hospital Geral de Nova lguaçu). Participants were enrolled at CUIMC beginning April 21^st^, 2020, and at the three clinical sites in Rio de Janeiro beginning August 15^th^, 2020. The trial was conducted in accordance with Good Clinical Practice guidelines, the Declaration of Helsinki, and the Brazilian National Ethics Committee Resolution 466/12. Written informed consent was obtained from all participants or from their legally authorized representative. The trial protocol was approved by institutional review boards at CUIMC and at each site in Rio de Janeiro (14) and is registered at ClinicalTrials.gov (Identifier: NCT04359810).

### Participants

Eligible participants were hospitalized patients aged ≥18 years with evidence of SARS-CoV-2 infection by polymerase chain reaction (PCR) of nasopharyngeal, oropharyngeal swab or tracheal aspirate sample within 14 days of randomization, with infiltrates on chest imaging and oxygen saturation ≤ 94% on room air or requirement for supplemental oxygen (including non-invasive positive pressure ventilation or high flow supplemental oxygen), IMV, or extracorporeal membrane oxygenation (ECMO) at the time of screening. Exclusion criteria included: participation in another clinical trial of anti-viral agent(s) for COVID-19; receipt of any anti-viral agent with possible activity against SARS-CoV-2 within 24 hours of randomization; duration of IMV or ECMO ≥ 5 days at time of screening; severe multi-organ failure; and a history of prior reactions to transfusion blood products. Following the U.S. Food and Drug Administration Emergency Use Authorization on May 1^st^, 2020 (15), concomitant use of remdesivir was permitted. The use of other treatments, including corticosteroids, was at the discretion of treating clinicians, and supportive care was provided according to standards at each site.

### Procedures

Convalescent plasma used at all study sites was collected by the New York Blood Center from patients who had recovered from laboratory-confirmed COVID-19, provided informed consent, had a minimum anti-SARS-CoV-2 total IgG antibody titer of ≥1:400 by quantitative enzyme linked immunosorbent assay against the spike protein (16), were at least 14 days asymptomatic following resolution of COVID-19, and had a negative PCR test for SARS-CoV-2 from a nasopharyngeal swab. Control plasma consisted of oldest available plasma at each study site without prior testing for anti-SARS-CoV-2 antibodies; all control plasma was collected prior to January 1^st^, 2020 in Rio de Janeiro and February 20^th^, 2020 in New York City. For all participants who received their treatment assignment, a single unit of plasma (~200-250 milliliters) was transfused over approximately 2 hours. Titers of neutralizing anti-SARS-CoV-2 antibody were measured in convalescent plasma units post hoc. Neutralization titer was determined with a SARS-CoV-2 viral neutralization assay which measured inhibition of virus growth after exposure to serial plasma dilutions using quantitative real-time reverse transcription-PCR (qRT-PCR). Further details are described in the protocol (14) and supplement. Given concern for emerging viral variants, we performed genomic sequencing of SARS-CoV-2 on nasopharyngeal swab samples from a subset of patients enrolled in Brazil. Sequences were mapped to the SARS-CoV-2 reference genome (sequence NC_045512) in NCBI. Additional details are included in the supplement.

### Randomization and Blinding

Enrolled participants were randomized in a 2:1 ratio to receive either convalescent plasma or control plasma using a web-based randomization platform; treatment assignments were generated using randomly permuted blocks of different sizes. Participants were transfused within 48 hours of randomization. The clinical teams directly managing patients and the trial clinicians who adjudicated clinical status and determined 28-day outcomes were blinded to treatment allocation. The hospital blood bank at each site and the clinical research teams who completed case record forms and performed other study specific procedures were not blinded; this was done to prevent errors in treatment allocation.

### Outcomes

The primary outcome was clinical status at day 28 following randomization, measured using an ordinal scale based on that recommended by the World Health Organization (17): 1, not hospitalized with resumption of normal activities; 2, not hospitalized, but unable to resume normal activities; 3, hospitalized, not requiring supplemental oxygen; 4, hospitalized, requiring supplemental oxygen; 5, hospitalized, requiring high-flow oxygen therapy or noninvasive mechanical ventilation; 6, hospitalized, requiring ECMO, IMV, or both; 7, death. Since distinguishing between clinical status 1 and 2 on the ordinal scale was difficult in participants discharged from hospital, these two scores were combined, and a six-point ordinal scale was used for all analyses of the primary outcome. Pre-specified secondary outcomes included time-to-clinical improvement (defined as improvement in at least one point from baseline on the ordinal scale or alive at discharge from hospital, whichever came first), in-hospital mortality, 28-day mortality, time-to-discontinuation of supplemental oxygen, time-to-hospital discharge, and serious and grade 3 and 4 adverse events.

The initial primary outcome was time-to-clinical-improvement. However, it became clear that this primary outcome would not reflect instances when patients’ clinical status subsequently worsened after improvement. Thus, the primary outcome of the study was amended to clinical status at day 28, and time-to-clinical-improvement became a secondary outcome. This change was made on August 8^th^, 2020 (at which point 31% [70/223] of the trial population was enrolled) without any knowledge of outcome data, and the protocol was updated accordingly with approval of the data safety and monitoring board (14).

Clinical status and adverse events were assessed daily during hospitalization through review of medical records and/or in-person visits. For participants discharged prior to day 28, clinical status and adverse events were determined via telephone and/or in-person visits. In patients who were discharged from hospital alive and not reachable for day 28 assessment, the last available clinical status was carried forward for the primary analysis, and sensitivity analyses were performed to account for potential bias due to loss-to-follow-up.

### Statistical Analysis

The trial was analyzed by comparing patients randomized to convalescent plasma versus control plasma, with patients randomized to control plasma serving as the reference group. The primary outcome was analyzed using a one-sided Mann-Whitney test for an alternative hypothesis favoring the convalescent plasma arm (a “go” decision in this phase 2 trial). To assess the magnitude of clinical effects, an odds ratio (OR) for improved clinical status on the modified ordinal scale was estimated under the proportional odds model. An OR >1.0 indicated improved clinical status among patients randomized to convalescent plasma versus control plasma. Post-hoc analyses of the primary outcome were also performed using a multivariable proportional odds model including age, sex, and duration of illness at baseline, prognostic factors imbalanced between treatment groups after randomization. Additional details are available in the supplement.

Pre-specified subgroups in analyses of the primary outcome were defined according to level of respiratory support at randomization (no supplemental oxygen, supplemental oxygen [including high-flow oxygen therapy and noninvasive ventilation], IMV or ECMO) and symptom duration at randomization (≤7□days, >□7□days) (14). Post hoc subgroup analyses were performed according to study country, age, sex, concomitant treatment with corticosteroids, and by titers of neutralizing anti-SARS-CoV-2 antibody in infused convalescent plasma units.

For the initial primary outcome of time-to-clinical-improvement, the intended sample size was 129 participants. However, after the primary outcome was amended, the sample size was re-calculated based on blinded pooled data of day 28 outcomes from an interim analysis by the data safety and monitoring board (July 2^nd^, 2020) and an OR of 1.7 under a proportional odds assumption. With a 2:1 randomization ratio and a total sample size of 219 participants (146 in the convalescent plasma arm versus 73 in the control arm), we determined that a one-sided Mann-Whitney test at a level of 15% would have 82% power to detect an OR 1.7. At the time the primary outcome was amended, a recent trial of remdesivir reported an OR 1.50 with 95% confidence interval (CI) of 1.18–1.91, which overlapped with our assumed OR (18).

Between group differences are reported using point estimates (OR or hazard ratio [HR]), with 95% confidence intervals and p-values. The p-value for the Mann-Whitney test in the primary outcome analysis (“go vs. “no-go” decision) is one-sided. All other p-values including those associated with point estimates are 2-sided and without adjustment for multiple comparisons. Analyses were performed using SAS, version 9.4 (SAS Institute, Cary, NC, USA).

## Results

### Participants

Between April 21^st^ and November 27^th^, 2020, a total of 630 patients were evaluated for inclusion criteria across the five study sites. Two-hundred-twenty-three were enrolled, randomized and included in the intention to treat (ITT) analysis (**Figure 1**). Four participants were randomized but did not receive their assigned treatment: three participants (two randomized to convalescent plasma and one to control plasma) had improvements in oxygen saturation to >94% prior to transfusion, and one participant randomized to convalescent plasma developed a maculopapular rash prior to receipt of plasma for which subsequent transfusion was deferred.

**Figure 1:**
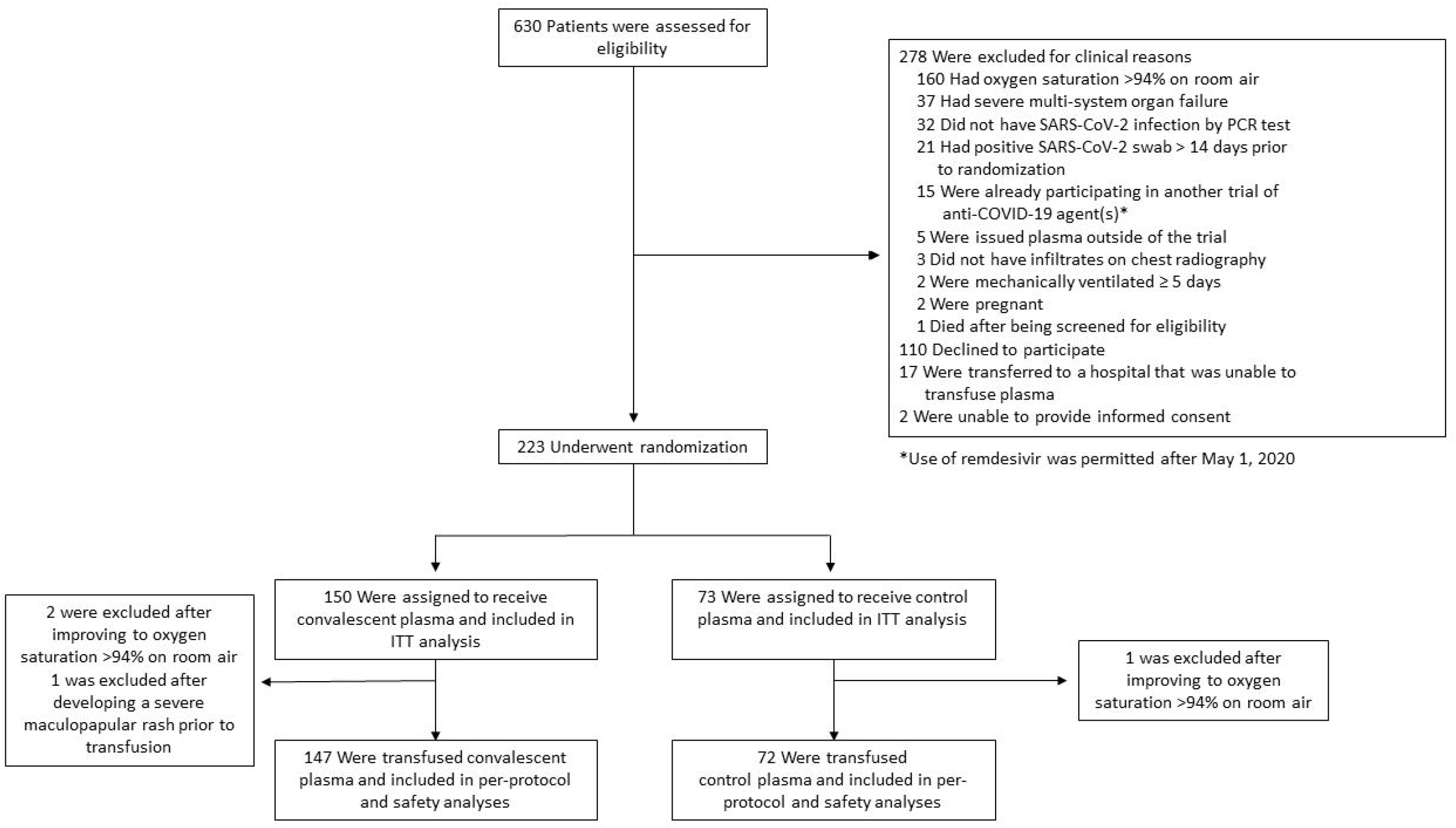
Trial flow diagram

Thus, 219 patients were included in the per-protocol and safety analysis: 147 participants transfused convalescent plasma, and 72 participants transfused control plasma (**Figure 1**). Data on neutralizing antibody titers were available for 89% (130/150) of convalescent plasma units. Of these, the median titer was 1:160 (IQR 1:80-1:320).

Of the 223 participants enrolled, 73 were enrolled in New York City and 150 in Rio de Janeiro (**Table 1**). The median age of participants was 61 years and 66% (147/223) were male. The median duration of symptoms prior to randomization was 9 days. Nearly all participants required respiratory support at baseline: 57% (126/223) of participants required supplemental oxygen, 25% (55/223) required high-flow oxygen therapy or non-invasive mechanical ventilation, and 13% (28/223) required IMV or ECMO. Some imbalances were present between treatment groups; participants enrolled in the convalescent plasma group were younger, with fewer men and a slightly longer symptom duration. During the trial period, 81% (181/223) of participants received corticosteroids and 6% (13/223) received remdesivir, the latter exclusively in New York City.

**Table 1:** Baseline patient characteristics.

| Variable | Convalescent<br>plasma<br>N = 150 | Normal control<br>plasma<br>N = 73 |
| --- | --- | --- |
| **Sex, n (%)** |  |  |
| Male | 96 (64) | 51 (70) |
| Female | 54 (36) | 22 (30) |
| **Age in years, median (IQR)** | 60 (48-71) | 63 (49-72) |
| **Age group, n (%)** |  |  |
| <60 years | 74 (49) | 28 (38) |
| 60-69 years | 35 (23) | 24 (33) |
| 70-79 years | 28 (19) | 16 (22) |
| ≥80 years | 13 (9) | 5 (7) |
| **Geographic location** |  |  |
| United States | 49 (33) | 24 (33) |
| Brazil | 101 (67) | 49 (67) |
| **Body mass index<sup>a</sup>** |  |  |
| BMI, median (IQR) | 30.1 (26.6-34.7) | 29.4 (26.2-33.0) |
| BMI ≥ 30 kg/m<sup>2</sup> | 76 (51) | 33 (45) |
| **Baseline conditions, n (%)** |  |  |
| Hypertension | 53 (35) | 22 (30) |
| Diabetes mellitus | 55 (37) | 27 (37) |
| Chronic cardiac disease | 56 (37) | 28 (38) |
| Chronic kidney disease | 13 (9) | 8 (11) |
| Chronic pulmonary disease | 15 (10) | 5 (7) |
| Chronic liver disease | 3 (2) | 1 (1) |
| HIV | 4 (3) | 0 (0) |
| Hyperlipidemia | 27 (18) | 9 (12) |
| **Duration of COVID-19**<br>**symptoms prior to**<br>**randomization, days, median**<br>**(IQR)<sup>b</sup>** | 10 (7-13) | 9 (7-11) |
| **Symptoms reported, n (%)** |  |  |
| Shortness of breath | 125 (83) | 58 (79) |
| Fever | 66 (44) | 27 (37) |
| Cough | 114 (76) | 49 (67) |
| **Clinical status at randomization**<br>**based on ordinal scale<sup>c</sup>** |  |  |
| 3: Hospitalized, not requiring<br>supplemental oxygen | 5 (3) | 5 (7) |
| 4-5: Hospitalized, requiring<br>supplemental oxygen, HFO, NIV | 125 (83) | 57 (78) |
| 6: Hospitalized, requiring IMV,<br>ECMO, or both | 17 (11) | 11 (15) |
| **Concomitant medications**<br>**received during study period** |  |  |
| Corticosteroids | 121 (81) | 60 (82) |
| Remdesivir | 8 (5) | 5 (7) |
| Hydroxychloroquine | 8 (5) | 5 (7) |
| Antibacterial agent | 111 (74) | 60 (82) |

Abbreviations: BMI: body mass index, IQR: interquartile range, HFO: high-flow oxygen therapy, NIV: non-invasive mechanical ventilation, IMV: invasive mechanical ventilation, ECMO: extracorporeal membrane oxygenation
Legend: <sup>a</sup>Unknown for 4 patients (2 in each treatment group), <sup>b</sup>Unknown for 6 patients (3 in each treatment group), <sup>c</sup>Baseline outcome assessment unknown for 3 patients (all in convalescent plasma group).

Primary outcome assessment of clinical status at 28 days was completed for 215 (96%) of 223 randomized patients. Eight participants with indeterminate clinical status at day 28 were discharged alive but were unable to be contacted at day 28. Of these eight participants, three had ≥14 days of follow-up and five had <14 days of follow-up.

### Primary outcome

Although participants randomized to receive convalescent plasma had 1.5 times the odds of a one-point improvement in clinical status at day 28, this difference was not statistically significant (OR 1.5, 95% confidence interval [CI] 0.83-2.68, p=0.18 (**Table 2**)). After adjustment for age, sex, and illness duration, the odds of improvement were similar (**Table 2**). Results were also similar in unadjusted and adjusted analyses of the per-protocol population and in two sensitivity analyses, one in which the 8 participants without a definitive day 28 outcome were considered deceased, and another in which the last available clinical status was carried forward for patients with ≥14 days of follow-up and patients with <14 days of follow-up were considered deceased (**Tables S1-S3**). Using a one-sided Mann-Whitney test of the alternative hypothesis favoring the convalescent plasma arm, the primary outcome analysis of the ITT population was consistent with a “go” decision (p=0.09) in this phase 2 clinical trial.

**Table 2:** Clinical efficacy outcomes among patients randomized to convalescent plasma versus control plasma (intention-to-treat population)

| Outcomes | Convalescent Plasma<br>N=150 | Control Plasma<br>N=73 | OR or sHR<br>(95% CI) | P-value | Adjusted<sup>a</sup> OR<br>or sHR<br>(95% CI) | P-value |
| --- | --- | --- | --- | --- | --- | --- |
| **Primary outcome, clinical status at 28-days, n (%)** |  |  | OR 1.50<br>(0.83-2.68) | 0.180 | OR 1.38<br>(0.73-2.61) | 0.318 |
| 1 and 2: Not hospitalized | 108 (72.0) | 48 (65.8) |  |  |  |  |
| 3: Hospitalized, not requiring supplemental oxygen | 3 (2.0) | 2 (2.7) |  |  |  |  |
| 4: Hospitalized, requiring supplemental oxygen | 7 (4.7) | 1 (1.4) |  |  |  |  |
| 5: Hospitalized, requiring high-flow oxygen therapy or noninvasive mechanical ventilation | 1 (0.7) | 0 (0.0) |  |  |  |  |
| 6: Hospitalized, requiring IMV, ECMO, or both | 12 (8.0) | 4 (5.5) |  |  |  |  |
| 7: Dead | 19 (12.6) | 18 (24.6) |  |  |  |  |
| **Secondary outcomes** |  |  |  |  |  |  |
| Time-to-clinical improvement, median,<sup>b</sup> days (IQR) | 5 (4-6) | 7 (5-8) | sHR 1.21<br>(0.89-1.65) | 0.231 | sHR 1.20<br>(0.87-1.64) | 0.261 |
| In-hospital mortality,<sup>c</sup> n (%) | 19 (12.6) | 18 (24.6) | OR 0.44<br>(0.22-0.91) | 0.034 | OR 0.47<br>(0.21-1.06) | 0.068 |
| 28-day mortality,<sup>c</sup> n (%) | 19 (12.6) | 18 (24.6) | OR 0.44<br>(0.22-0.91) | 0.034 | OR 0.47<br>(0.21-1.06) | 0.068 |
| Time-to-discontinuation of supplemental oxygen,<sup>d</sup> median, days (IQR) | 6 (3-16) | 7 (3-11) | sHR 1.12<br>(0.80-1.56) | 0.508 | sHR 1.12<br>(0.80-1.56) | 0.514 |
| Time-to-hospital-discharge, median, days (IQR) | 9 (6-28) | 8 (6-22) | sHR 1.05<br>(0.77-1.43) | 0.756 | sHR 1.02<br>(0.75-1.38) | 0.913 |

Abbreviations: CI: confidence interval, ECMO: extracorporeal membrane oxygenation, sHR: subhazard ratio, IMV: invasive mechanical ventilation, IQR: interquartile range, OR: odds ratio.
Legend: <sup>a</sup>Adjusted for age (continuous variable), sex, and duration of symptoms at baseline (duration of symptoms unknown for 6 patients); <sup>b</sup>Baseline outcome assessment unknown for 3 patients (all in treatment group); <sup>c</sup>No patients were known to have died following discharge from hospital; <sup>d</sup>13 patients excluded from unadjusted analysis (10 participants enrolled but did not require supplemental oxygen, 3 patients without a baseline assessment; 16 patients excluded from adjusted analysis).

### 28-day Mortality

In the ITT population, mortality at 28 days was significantly lower among participants randomized to convalescent versus control plasma (19/150 [12.6%] versus 18/73 [24.6%], OR 0.44, 95% CI 0.22-0.91, p=0.034) when the last available clinical status was carried forward for the 8 patients without definitive day 28 outcome status (**Table 2, Figure 2**). These results were consistent in adjusted analyses and in sensitivity analyses to account for the 8 patients without definitive day 28 outcome (**Tables 2 and S4-S5**). All recorded deaths occurred during hospitalization. No significant between-group differences were observed in the other secondary outcomes (**Table 2 and Figure 3**).

**Figure 2:**
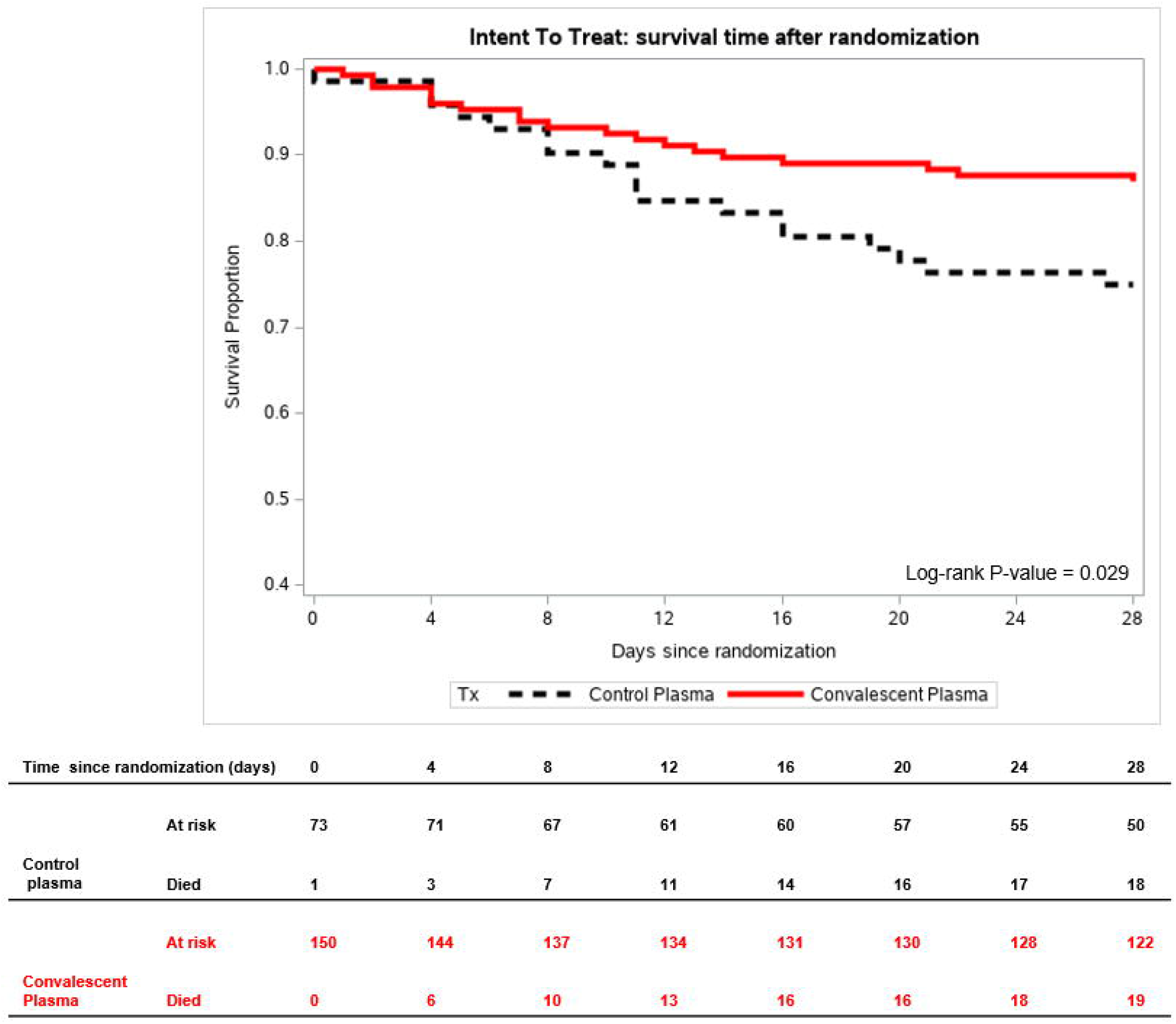
Kaplan-Meier estimates of mortality, stratified by treatment group.

**Figure 3:**
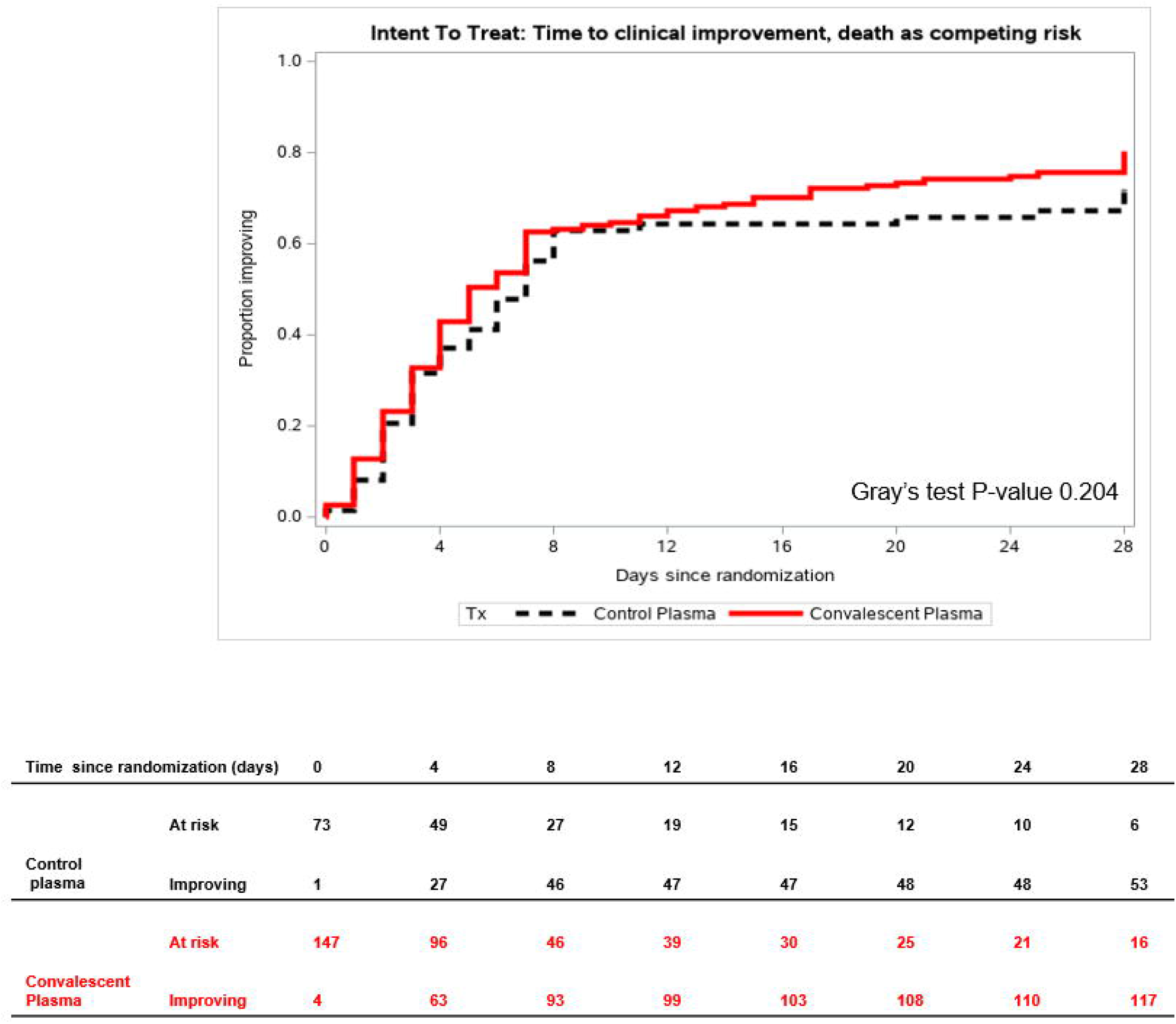
Time-to-clinical improvement with death considered a competing risk, stratified by treatment group.

### Subgroup Analyses

In pre-specified analyses of the primary outcome based on respiratory support and symptom duration at baseline, no significant between-group differences were observed in the primary outcome (**Figure 4 and Tables S6-S7**). However, we observed trends towards improved clinical status among patients who received convalescent plasma ≤7 days after symptom onset and those who received convalescent plasma with higher-titers of neutralizing antibody and concomitant corticosteroids (**Figures 4 and S1-S2 and Tables S6-S7**). In stratified analyses of 28-day mortality, unadjusted point-estimates consistently favored the convalescent plasma group **(Figure S3)**.

**Figure 4:**
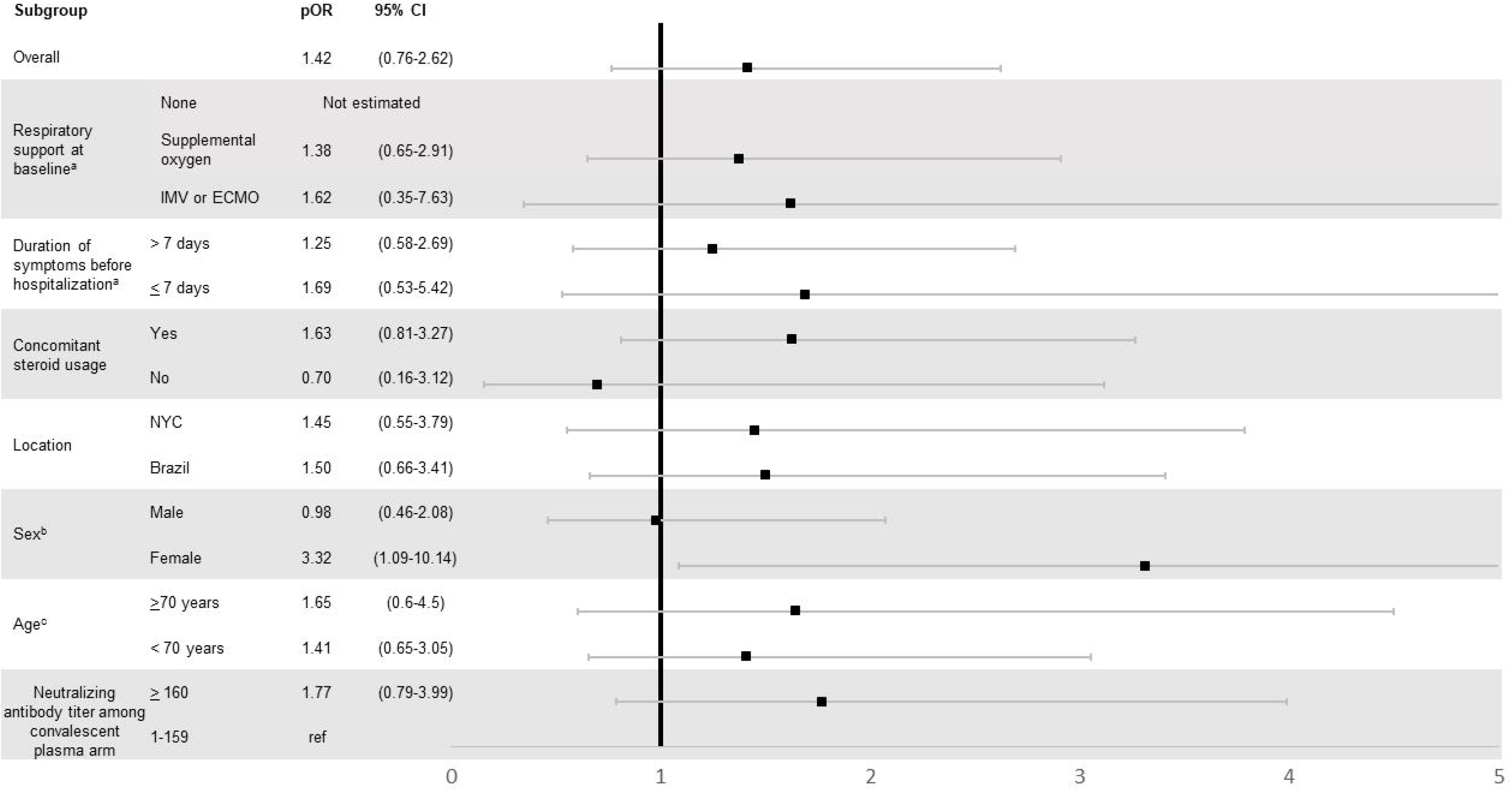
Subgroup analyses of primary outcome of clinical status at 28 days, adjusted for age and sex

### SARS-CoV-2 genomic sequencing

RNA template was sufficient to recover near complete (>99%) genomic sequence from 40 nasopharyngeal samples from Brazil. Twenty-nine (73%) represented common clades circulating worldwide and had no spike protein mutations. None of the samples contained the mutations characteristic of B.1.1.28 P1. Four had mutations found in B.1.1.28 (E484K) but did not have the N501Y, K417N/T mutations found in P1. One sample had 3 of 4 mutations characteristic of B.1.1.28 (AM-II), including V1176K in S, that is not known to impair neutralization. In short, we found no evidence of neutralization-escape mutants.

### Safety Analysis

Serious adverse events occurred in 39 of 147 (26.5%) patients who received convalescent plasma and 26 of 72 (36.1%) patients who received control plasma (**Tables S8-S11**). Adverse events considered as definitely or probably associated with plasma transfusion were reported in 4 of 147 (2.7%) patients who received convalescent plasma and 3 of 72 (4.2%) patients who received control plasma. In patients who received convalescent plasma, these events included worsening anemia, urticaria, skin rash, and transfusion-associated circulatory overload.

## Discussion

In this randomized, blinded, and controlled phase 2 trial conducted in New York City and Rio de Janeiro, treatment with convalescent plasma as compared to control plasma did not result in significant clinical improvement at 28 days, based on an ordinal scale of clinical status, among adults hospitalized with severe and critical COVID-19. However, mortality at 28 days was significantly lower among patients randomized to convalescent plasma. This effect on mortality was observed across analyses adjusted for imbalances in baseline variables with prognostic relevance and in sensitivity analyses performed to account for indeterminate 28-day vital status in 8 patients.

Although limited, available data suggest that treatment efficacy for convalescent plasma may be dependent on illness duration and severity and titers of neutralizing anti-SARS-CoV-2 antibody in transfused plasma. In a recent clinical trial from Argentina, transfusion of high-titer convalescent plasma within 72 hours of symptom onset prevented progression to severe illness among elderly adults with mild COVID-19 (19). In contrast, no overall improvements in clinical status were observed in recent trials of convalescent plasma among inpatients with severe COVID-19 in China and Argentina (11,13). However, subgroup analyses in these trials suggested a possible benefit among patients with less severe and shorter durations of illness. These signals are consistent with results of a retrospective study of over 3,000 U.S. adults who received convalescent plasma for treatment of severe COVID-19 (10). In this retrospective analysis, high-titer convalescent plasma was associated with improved mortality among inpatients who were not receiving IMV at the time of transfusion. Considering power limitations of our trial, we observed similar trends towards improvement in the primary outcome among patients in the convalescent plasma group transfused within 7 days of symptom onset and those who received convalescent plasma with higher-titers of neutralizing anti-SARS-CoV-2 antibody.

In the context of emerging SARS-CoV-2 variants, some of which may be associated with greater transmissibility and more severe illness (20), convalescent plasma may offer distinct therapeutic advantages. Since convalescent plasma, which contains polyclonal antibodies, may be donated and transfused locally, and its use may be more adaptable to rapidly changing local viral ecology than other interventions. In contrast, monoclonal antibody therapies may need to be repeatedly engineered and combined to optimize potency among emergent SARS-CoV-2 variants (21,22). Further, since collection and distribution of convalescent plasma units can be performed using existing blood donation protocols and infrastructure, convalescent plasma may be more scalable for use in low- and middle-income countries.

Although clinical status at 28 days was not significantly different between treatment groups, 28-day mortality was significantly lower among patients randomized to receive convalescent plasma. Although this secondary outcome was pre-specified, our study was not powered to detect a difference in mortality and analyses of our secondary outcomes were not adjusted for multiplicity. This finding should be interpreted with caution until full results from larger inpatient trials adequately powered to detect differences in mortality, such as the Randomised Evaluation of COVID-19 Therapy (RECOVERY), are available (23).

We observed no significant difference in adverse events between treatment groups and very few events were considered related to plasma infusion. Although use of control plasma may have potentially contributed to hypercoagulability (24), the incidence of thrombotic events in our study population was similar to that reported in observational studies of patients with severe COVID-19 (25).

Our trial has several strengths. First, the randomized, blinded, controlled design of our trial was implemented with high adherence to the study protocol. Second, we enrolled severe and critical COVID-19 patients in racially and ethnically diverse urban settings in two countries. Third, our strategy for qualification and collection of convalescent plasma was pragmatic, increasing generalizability of our findings to settings where quantification of neutralization activity is unavailable. However, we quantified neutralizing antibody titers in approximately 90% of convalescent plasma samples post hoc. Fourth, our use of control plasma was a significant strength since both study agents had the same appearance, enhancing the blinded nature of the trial, and both had a similar effect on volume expansion. As convalescent plasma may have other immunomodulatory factors apart from anti-SARS-CoV-2 antibodies, such as immunoglobulins, hemostatic proteins and cytokines, use of normal plasma as a comparator allowed us to evaluate the effect of convalescent antibodies while controlling for these other factors.

Our trial has several limitations. First, although convalescent plasma was collected from donors with anti-SARS-CoV-2 total IgG antibody titer of ≥1:400, neutralizing antibody titers in some convalescent plasma units were low, and we do not have data on antibody titers in patient samples pre- and post-transfusion. Second, while all control plasma units were collected prior to the first known cases of COVID-19 in Rio de Janeiro and New York City, one out of 19 units tested neutralized SARS-CoV-2 at low titer. Although this could represent a false-positive, it is possible that other control plasma units could have contained anti-coronavirus antibodies. Third, supportive care was not standardized across study sites. However, we observed no significant differences in outcomes in stratified by country. Fourth, the trial was underpowered for secondary outcomes and multiple comparisons.

In conclusion, although the use of convalescent plasma was not associated with improved day 28 clinical status based on an ordinal scale, it was associated with a significant reduction in day 28 mortality. This result should be interpreted with caution pending results from larger inpatient trials and may warrant further evaluation.

## Data Availability

Data will only be available as per the terms of the informed consent and institutional policies around data transfer.

## Contributors

MRO’D and WIL conceived the study and led protocol development. MRO’D, MJC, JJ, NMP, AE, KC, and WIL contributed to study design. MRO’D, BG, MJC, CME, NMP, MRL, YKC, EJ, JHP, MPD, SWC, DA, KR, LCS, AV, VGV, DS, BJM, SDJ, and WIL contributed to data acquisition, analysis, and/or interpretation. NM, LC, TB, and WIL performed and interpreted neutralization assay experiments. EH, ZCB, SLS, ES, FDZ, FLC, KEH, SAF, JS, BS, WHL, SW, and BS contributed to convalescent plasma collection, qualification, and release. SB, AK, AW, and NMP coordinated study activities. MRL and KC performed statistical analyses. MRO’D, MJC, MRL, YKC, VG, and WIL wrote the manuscript. All authors contributed to critical revision of the manuscript. MRO’D, MRL, NMP, and KC had full access to the data, and take responsibility for the integrity of the data and the accuracy of the analysis.

## Data Sharing and Reproducible Research

The study protocol, definition of outcomes, and other relevant materials have been published previously (14). De-identified participant data will be made available to researchers affiliated with an appropriate institution within 3 months of publication following mutual signing of a data access agreement and obtainment of necessary ethics approvals. The trial investigators must approve all proposals to access the data and have the right to review and comment on any draft manuscripts before publication. Requests for data access should be sent to Dr. Max R. O’Donnell. Statistical code used in study analyses is also available on request.

## Acknowledgements

This trial was funded by an unrestricted grant from the Amazon Foundation to Columbia University. The authors would like to thank the patients who participated in this study and their families as well as the members of the data safety and monitoring board (Neil W. Schluger, Scott M. Hammer, Deborah Donnell).

