## Supplement for "A randomized, double-blind, controlled trial of convalescent plasma in adults with severe COVID-19"

**Supplementary Material**

**Supplemental Methods**

Collection and qualification of convalescent plasma units

*Donor criteria*

Inclusion criteria for individuals donating convalescent plasma were: (1) prior laboratory confirmed SARS-CoV-2 or documented physician diagnosis; (2) at least 18 years of age; (3) weight >110 pounds; (4) subjective good health; (5) reported resolution of COVID-19 related symptom at least 14 days prior to enrollment; (6) fulfillment of New York Blood Center criteria for blood donation (https://www.nybc.org/donate-blood/become-donor/can-i-donate-blood/); (7) hemoglobin > 12.5 g/dL for females and >13.0 g/dL for males; (8) negative SARS-CoV-2 test on nasopharyngeal swab using an EUA-approved PCR-based assay; and (9) positive anti-SARS-CoV-2 antibody test against the spike trimer at 1:400 dilution as described below. Exclusion criteria included all the standard exclusions required for blood donation as per New York Blood Center Criteria (https://www.nybc.org/donate-blood/become-donor/can-i-donate-blood/). Each convalescent plasma unit was derived from a single donor, each of whom donated between one to four units by apheresis.

*Antibody Measurement*

The SARS-CoV-2 spike trimer was coated on 96-well ELISA plates at a concentration of 200 ng/well at 4°C overnight and unbound proteins were then removed by washing with PBS, followed by blocking with PBS/3% nonfat dry milk, as described (16). Plasma samples were serially diluted in PBS with Tween 20 (PBST; 0.1% Tween-20 in PBS) + 10% fetal calf serum (FCS) at 1:400 replicate dilutions into wells of the coated plate, which was incubated at 37 °C for 1 h, followed by washing 6 times with PBST. Peroxidase AffiniPure goat anti-human IgG (H+L) antibody (1:3,000 dilution; Thermo Fisher Scientific) was subsequently added to each well and incubated for 1 h at 37 °C, washed and tetramethylbenzidine substrate (Sigma-Aldrich) was added and the reaction was stopped using 1 M of sulfuric acid. Absorbance was measured at 450 nm and expressed as an optical density or OD_450_ value. Prior validation studies established a cutoff OD_450_ value for positivity, resulting in >99% specificity in samples obtained from the pre-COVID-19 era. This assay was approved for use by the New York State Department of Health prior to the availability of commercial anti-SARS-CoV-2 antibody assays.

Measurement of neutralizing anti-SARS-CoV-2 antibody titers

Virus neutralization tests (NT) were performed using SARS-CoV-2 strain 2019-nCoV/USA-WA1-2020 (provided by Rocky Mountain Laboratories, MT, USA). Test plasma was heat inactivated (30 min, 56 ^o^C) before being two-fold serially diluted in Dulbecco’s Modified Eagle Media (DMEM), starting at a 1:20 dilution. The diluted plasma (50 microliter) was mixed with virus (100 TCID_50_, 50 microliter) and incubated for 1 hour at 37^o^C. The plasma-virus mix was then added to a Vero E6 cell monolayer (ATCC CRL-1586; Multiplicity of Infection (MOI) = 0.01) and incubated (1.5 hours, 37^o^C, 5% CO_2_), before the inoculum was replaced by DMEM supplemented with 1% fetal calf serum. Culture supernatant was removed after 48 hours of incubation (37^o^C, 5% CO_2_) and virus growth determined using quantitative real-time reverse transcription-polymerase chain reaction (Triplex CII-SARS-CoV-2 rRT-PCR Test, EUA200510). The highest plasma dilution that prevented virus growth (cycle threshold [ct] <30) was rated as neutralization titer.

SARS-COV-2 genomic sequencing

Extracts of nasopharyngeal swab samples positive for SARS-CoV-2 RNA using the Triplex CII-SARS-CoV-2 rRT-PCR assay were reverse transcribed for Illumina sequencing using the Kapa HyperPlus kit (Roche) employing enzymatic fragmentation to about 300 basepair average size. Prior to sequencing (Ilumina NextSeq 550, 150 cycles single-read sequencing) SARS-CoV-2 sequences were enriched through hybridization to biotinylated myBaits Expert Virus-SARS-CoV-2 baits (Daicel Arbor Biosciences). The biotinylated oligonucleotides were captured by streptavidin beads, washed, and amplified through 14 cycles of PCR with Illumina sequencing primers. Raw reads (5-28 million/barcoded sample) were quality filtered and mapped to the NCBI SARS-CoV-2 reference sequence NC_045512 (6-11 million mapped reads per sample).

Statistical Analysis

The trial was analyzed by comparing patients randomized to convalescent plasma versus control plasma, with patients randomized to control plasma serving as the reference group. The primary outcome was analyzed using a one-sided Mann-Whitney test for an alternative hypothesis favoring the convalescent plasma arm (a “go” decision in this proof-of-concept phase 2 trial). To assess the magnitude of clinical effects, an odds ratio for improved clinical status on the modified ordinal scale was estimated under the proportional odds model. An odds ratio >1.0 would indicate improved clinical status on the ordinal scale among patients randomized to convalescent plasma versus control plasma. Post-hoc analyses of the primary outcome were also performed using a multivariable proportional odds model including age, sex, and duration of illness at baseline, prognostic factors imbalanced between treatment groups after randomization.

To evaluate the association between treatment group and time-to-clinical improvement, time-to-discontinuation of supplemental oxygen, and time-to-hospital discharge, we used Fine and Gray regression models (considering death as a competing risk) to estimate sub-hazard ratios and the cumulative incidence of clinical improvement as a function of time. For analyses of in-hospital and 28-day mortality, we estimated odds ratios using logistic regression models and estimated survival at 28 days using the Kaplan-Meier method. Fine and Gray and logistic regression models were performed both unadjusted and adjusted for age, sex, and duration of illness.

Sensitivity analyses for the primary outcome and mortality at 28-days included the per-protocol population and populations in which patients without definitive determination of clinical status at 28-days (those whose clinical status was carried forward in the primary outcome analysis) were treated as deceased, and another in which the last available clinical status was carried forward for patients with ≥14 days of follow-up, and patients with <14 days of follow-up were considered deceased.

Pre-specified subgroups in analyses of the primary outcome were defined according to level of respiratory support at randomization (no supplemental oxygen, supplemental oxygen [including high-flow oxygen therapy and noninvasive ventilation], IMV or ECMO) and symptom duration at randomization (≤7 days, > 7 days) (14). Post hoc subgroup analyses were also performed according to study site (New York City vs. Rio de Janeiro), age, sex, concomitant treatment with corticosteroids, and by titers of neutralizing anti-SARS-CoV-2 antibody in infused convalescent plasma units.

For the initial primary outcome of time-to-clinical-improvement, the intended sample size was 129 participants. However, after the primary outcome was amended to clinical status at day 28, the sample size was re-calculated. This calculation was based on blinded pooled data of day 28 outcomes from an interim analysis by the data safety and monitoring board (July 2^nd^, 2020) and an odds ratio of 1.7 under a proportional odds assumption. With a 2:1 randomization ratio and a total sample size of 219 participants (146 in the convalescent plasma arm versus 73 in the control arm), we determined that a one-sided Mann-Whitney test at a level of 15% would have 82% power to detect an odds ratio of 1.7. At the time the primary outcome was amended, a recent trial of remdesivir reported an odds ratio of 1.50 with 95% confidence interval (CI) of 1.18–1.91 (18). As this overlapped our assumed odds ratio, we increased enrollment to a total of 219 participants across all clinical sites.

Continuous variables are reported as medians (interquartile ranges) and categorical variables are summarized as counts and percentages. Between group differences are reported using point estimates (odds ratio or hazard ratio), with 95% confidence intervals and p-values. The p-value for the Mann-Whitney test in the primary outcome analysis (“go vs. “no-go” decision) is one-sided. All other p-values including those associated with point estimates are 2-sided and without adjustment for multiple comparisons. Analyses were performed using SAS, version 9.4 (SAS Institute, Cary, NC, USA).

**Supplemental Results**

| **Primary outcome, clinical status at 28-days, n (%)** | **Convalescent Plasma**  **N=147** | **Control Plasma**  **N=72** | **OR (95% CI)** | **P-value** | **Adjusted^a^ OR (95% CI)** | **P-value** |
| --- | --- | --- | --- | --- | --- | --- |
| 1 and 2: Not hospitalized | 105 (71.4) | 47 (65.3) | 1.49 (0.83-2.68) | 0.19 | 1.43 (0.76-2.71) | 0.27 |
| 3: Hospitalized, not requiring supplemental oxygen | 3 (2.0) | 2 (2.8) |  |  |  |  |
| 4: Hospitalized, requiring supplemental oxygen | 7 (4.8) | 1 (1.4) |  |  |  |  |
| 5: Hospitalized, requiring high-flow oxygen therapy or noninvasive mechanical ventilation | 1 (0.7) | 0 (0.0) |  |  |  |  |
| 6: Hospitalized, requiring IMV, ECMO, or both | 12 (8.2) | 4 (5.6) |  |  |  |  |
| 7: Dead | 19 (12.9) | 18 (25.0) |  |  |  |  |

**Table S1: Primary outcome analysis among patients who received convalescent plasma versus control plasma (per-protocol population)**

Abbreviations: CI: confidence interval, ECMO: extracorporeal membrane oxygenation, HR: hazard ratio, IMV: invasive mechanical ventilation, IQR: interquartile range, OR: odds ratio.

Legend: ^a^Adjusted for age (continuous variable), sex, and duration of symptoms at baseline (symptom duration unknown for 6 patients).

| **Primary outcome, clinical status at 28-days, n (%)** | **Convalescent Plasma**  **N=150** | **Control Plasma**  **N=73** | **OR (95% CI)** | **P-value** | **Adjusted^a^ OR (95% CI)** | **P-value** |
| --- | --- | --- | --- | --- | --- | --- |
| 1 and 2: Not hospitalized | 102 (68) | 46 (63) | 1.38 (0.78-2.45) | 0.280 | 1.30 (0.70-2.39) | 0.404 |
| 3: Hospitalized, not requiring supplemental oxygen | 3 (2) | 2 (3) |  |  |  |  |
| 4: Hospitalized, requiring supplemental oxygen | 7 (5) | 1 (1) |  |  |  |  |
| 5: Hospitalized, requiring high-flow oxygen therapy or noninvasive mechanical ventilation | 1 (1) | 0 (0) |  |  |  |  |
| 6: Hospitalized, requiring IMV, ECMO, or both | 12 (8) | 4 (5) |  |  |  |  |
| 7: Dead | 25 (17) | 20 (27) |  |  |  |  |

**Table S2: Primary outcome analysis among patients randomized to convalescent plasma versus control plasma (intention-to-treat population), with 8 patients with clinical status carried forward considered deceased**

Abbreviations: CI: confidence interval, ECMO: extracorporeal membrane oxygenation, IMV: invasive mechanical ventilation

Legend: ^a^Adjusted for age (continuous variable), sex, and duration of symptoms at baseline (symptom duration unknown for 6 patients).

**Table S3: Primary outcome analysis among patients randomized to convalescent plasma versus control plasma (intention-to-treat population), with the last observation carried forward for 3 patients with ≥14 days of follow-up; remaining 5 with <14 days of follow-up considered deceased**

| **Primary outcome, clinical status at 28-days, n (%)** | **Convalescent Plasma**  **N=150** | **Control Plasma**  **N=73** | **OR (95% CI)** | **P-value** | **Adjusted^a^ OR (95% CI)** | **P-value** |
| --- | --- | --- | --- | --- | --- | --- |
| 1 and 2: Not hospitalized | 104 (69) | 47 (64) | 1.39 (0.78-2.47) | 0.280 | 1.31 (0.70-2.44) | 0.394 |
| 3: Hospitalized, not requiring supplemental oxygen | 3 (2) | 2 (3) |  |  |  |  |
| 4: Hospitalized, requiring supplemental oxygen | 7 (5) | 1 (1) |  |  |  |  |
| 5: Hospitalized, requiring high-flow oxygen therapy or noninvasive mechanical ventilation | 1 (1) | 0 (0) |  |  |  |  |
| 6: Hospitalized, requiring IMV, ECMO, or both | 12 (8) | 4 (5) |  |  |  |  |
| 7: Dead | 23 (15) | 19 (26) |  |  |  |  |

Abbreviations: CI: confidence interval, ECMO: extracorporeal membrane oxygenation, IMV: invasive mechanical ventilation

Legend: ^a^Adjusted for age (continuous variable), sex, and duration of symptoms at baseline (symptom duration unknown for 6 patients).

| **Vital status at day 28, n (%)** | **Convalescent Plasma**  **N=150** | **Control Plasma**  **N=73** | **OR (95% CI)** | **P-value** | **Adjusted^a^ OR (95% CI)** | **P-value** |
| --- | --- | --- | --- | --- | --- | --- |
| Dead | 25 (17) | 20 (27) | 0.53 (0.27-1.04) | 0.075 | 0.56 (0.27-1.17) | 0.120 |
| Alive | 125 (83) | 53 (73) |  |  |  |  |

**Table S4: Mortality at 28-days among patients randomized to convalescent plasma versus control plasma (intention-to-treat	population), with 8 patients with clinical status carried forward considered deceased**

Abbreviations: CI: confidence interval

Legend: ^a^Adjusted for age (continuous variable), sex, and duration of symptoms at baseline (symptom duration unknown for 6 patients).

**Table S5: Mortality at 28-days among patients randomized to convalescent plasma versus control plasma (intention-to-treat	population), with the last observation carried forward for 3 patients with ≥14 days of follow-up; remaining 5 with <14 days of	follow-up considered deceased**

Abbreviations: CI: confidence interval

Legend: ^a^Adjusted for age (continuous variable), sex, and duration of symptoms at baseline (symptom duration unknown for 6 patients).

| **Vital status at day 28, n (%)** | **Convalescent Plasma**  **N=150** | **Control Plasma**  **N=73** | **OR (95% CI)** | **P-value** | **Adjusted^a^ OR (95% CI)** | **P-value** |
| --- | --- | --- | --- | --- | --- | --- |
| Dead | 23 (15) | 19 (26) | 0.515 (0.26-1.02) | 0.068 | 0.53 (0.24-1.14) | 0.101 |
| Alive | 127 (85) | 54 (74) |  |  |  |  |

| **Primary outcome, clinical status at 28-days, n (%)** | **Convalescent Plasma** | | | **Control Plasma** | | | **Odds Ratio**  **(95% CI)** | | |
| --- | --- | --- | --- | --- | --- | --- | --- | --- | --- |
|  | IMV or ECMO  N=17 | Supplemental oxygen, HFO, NIV  N=125 | No supplemental oxygen  N=5 | IMV or ECMO  N=11 | Supplemental oxygen, HFO, NIV  N=57 | No supplemental oxygen  N=5 | IMV or ECMO | Supplemental oxygen, HFO, NIV | No supplemental oxygen |
| 1 and 2: Not hospitalized | 4 (23.5) | 98 (78.4) | 4 (80.0) | 2 (18.2) | 42 (73.7) | 4 (80.0) | 2.18 (0.51-9.29) | 1.38 (0.68-2.82) | 1.28 (0.06-27.91) |
| 3: Hospitalized, not requiring supplemental oxygen | 2 (11.8) | 0 (0.0) | 1 (20.0) | 1 (9.1) | 1 (1.8) | 0 (0.0) | **Adjusted^a^ Odds Ratio (95% CI)** | | |
| 4: Hospitalized, requiring supplemental oxygen | 1 (5.9) | 5 (4.0) | 0 (0.0) | 1 (9.1) | 0 (0.0) | 0 (0.0) | IMV or ECMO | Supplemental Oxygen, HFO, NIV | No supplemental oxygen |
| 5: Hospitalized, requiring high-flow oxygen therapy or noninvasive mechanical ventilation | 0 (0.0) | 1 (0.8) | 0 (0.0) | 0 (0.0) | 0 (0.0) | 0 (0.0) | 1.622 (0.35-7.63) | 1.38 (0.65-2.91) | Not calculable |
| 6: Hospitalized, requiring IMV, ECMO, or both | 4 (23.5) | 8 (6.4) | 0 (0.0) | 0 (0.0) | 4 (7.0) | 0 (0.0) |  |  |  |
| 7. Dead | 6 (35.3) | 13 (10.4) | 0 (0.0) | 7 (63.6) | 10 (17.5) | 1 (20.0) |  |  |  |

**Table S6: Primary outcome analysis among patients randomized to convalescent plasma versus control plasma (intention-to-treat population) according to level of respiratory support at baseline**

Abbreviations: CI: confidence interval, HFO: high-flow oxygen therapy, NIV: non-invasive mechanical ventilation, ECMO: extracorporeal membrane oxygenation, IMV: invasive mechanical ventilation

Legend: ^a^Adjusted for age and sex

| **Primary outcome, clinical status at 28-days, n (%)** | **Convalescent Plasma** | | **Control Plasma** | | **Odds Ratio**  **(95% CI)** | |
| --- | --- | --- | --- | --- | --- | --- |
|  | Symptoms >7 days  N=110 | Symptoms ≤7 days  N=37 | Symptoms >7 days  N=51 | Symptoms ≤7 days  N=19 | Symptoms >7 days | Symptoms ≤7 days |
| 1 and 2: Not hospitalized | 81 (73.6) | 26 (70.3) | 37 (72.5) | 10 (52.6) | 1.20 (0.58-2.49) | 1.92 (0.64-5.81) |
| 3: Hospitalized, not requiring supplemental oxygen | 3 (2.7) | 0 (0.0) | 1 (2.0) | 1 (5.3) | **Adjusted^a^ Odds Ratio (95% CI)** | |
| 4: Hospitalized, requiring supplemental oxygen | 5 (4.5) | 1 (2.7) | 1 (20.0) | 0 (0.0) | 1.25 (0.58-2.69) | 1.69 (0.53-5.42) |
| 5: Hospitalized, requiring high-flow oxygen therapy or noninvasive mechanical ventilation | 1 (0.9) | 0 (0.0) | 0 (0.0) | 0 (0.0) |  |  |
| 6: Hospitalized, requiring IMV, ECMO, or both | 8 (7.3) | 3 (8.1) | 1 (2.0) | 3 (15.8) |  |  |
| 7. Dead | 12 (10.9) | 7 (18.9) | 11 (21.6) | 5 (26.3) |  |  |

**Table S7: Primary outcome analysis among patients randomized to convalescent plasma versus control plasma (intention-to-treat population) according to duration of symptoms at baseline**

Abbreviations: CI: confidence interval, ECMO: extracorporeal membrane oxygenation, IMV: invasive mechanical ventilation

Legend: ^a^Adjusted for age and sex

**Table S8: All and transfusion-related adverse events in patients receiving**

**convalescent and control plasma**

| **Adverse event, n (%)** | **Convalescent Plasma**  **N=147** | **Control Plasma**  **N=72** |
| --- | --- | --- |
| Any adverse event | 96 (65.3) | 40 (55.6) |
| Grade 1 | 38 (25.9) | 17 (23.6) |
| Grade 2 | 61 (41.5) | 23 (31.9) |
| Grade 3 | 27 (18.4) | 17 (23.6) |
| Grade 4 | 26 (17.7) | 15 (20.8) |
| Death | 19 (12.9) | 18 (25.0) |
| Transfusion-related adverse event^a^ | 4 (2.7) | 3 (4.2) |

Legend: ^a^Considered 'definitely' or 'probably' associated with plasma infusion.

**Table S9: Description of transfusion-related adverse events**

| **Treatment Group** | **Description of event(s)** |
| --- | --- |
| Convalescent plasma: Patient A | Worsening anemia |
| Convalescent plasma: Patient B | Urticaria |
| Convalescent plasma: Patient C | Transfusion-associated circulatory overload  Hypersensitivity-attributed skin rash |
| Convalescent plasma: Patient D | Transfusion-associated circulatory overload |
| Control plasma: Patient E | Possible febrile non-hemolytic transfusion reaction  Urticaria |
| Control plasma: Patient F | Worsening anemia |
| Control plasma: Patient G | Transfusion-associated circulatory overload |

**Table S10: All adverse events, stratified by organ system**

| **All Adverse Events** | | **Convalescent Plasma**  **N=147** | | **Control Plasma**  **N=72** | |
| --- | --- | --- | --- | --- | --- |
|  |  | **N** | **%** | **N** | **%** |
| **Any Adverse Event** |  | 96 | 65.3 | 40 | 55.6 |
| Cardiovascular | any cardiovascular event | 39 | 26.5 | 24 | 33.3 |
|  | miscellaneous arrhythmia | 10 | 6.8 | 5 | 6.9 |
|  | atrial fibrillation | 3 | 2.0 | 1 | 1.4 |
|  | bradycardia | 6 | 4.1 | 4 | 5.6 |
|  | undifferentiated shock | 5 | 3.4 | 3 | 4.2 |
|  | cardiac arrest | 1 | 0.7 | 3 | 4.2 |
|  | congestive heart failure | 4 | 2.7 | 2 | 2.8 |
|  | extremity edema | 4 | 2.7 | 1 | 1.4 |
|  | elevated cardiac troponin | 1 | 0.7 | 0 | 0.0 |
|  | hypertension | 10 | 6.8 | 3 | 4.2 |
|  | hypotension | 8 | 5.4 | 6 | 8.3 |
|  | transfusion-associated circulatory overload | 2 | 1.4 | 1 | 1.4 |
|  | right ventricular dysfunction | 1 | 0.7 | 0 | 0.0 |
|  | septic shock | 4 | 2.7 | 4 | 5.6 |
|  | cardiogenic shock | 0 | 0.0 | 1 | 1.4 |
|  | myocarditis | 1 | 0.7 | 0 | 0.0 |
| Endocrine | any endocrine event | 9 | 6.1 | 4 | 5.6 |
|  | diabetic ketoacidosis | 2 | 1.4 | 0 | 0.0 |
|  | hyperglycemia | 4 | 2.7 | 2 | 2.8 |
|  | hypoglycemia | 3 | 2.0 | 1 | 1.4 |
|  | miscellaneous endocrine event | 0 | 0.0 | 2 | 2.8 |
| Gastrointestinal/hepatic | any gastrointestinal/hepatic event | 43 | 29.3 | 15 | 20.8 |
|  | abdominal pain | 4 | 2.7 | 3 | 4.2 |
|  | gastrointestinal bleeding | 0 | 0.0 | 3 | 4.2 |
|  | pneumoperitoneum | 1 | 0.7 | 0 | 0.0 |
|  | diarrhea | 9 | 6.1 | 3 | 4.2 |
|  | dysphagia | 2 | 1.4 | 1 | 1.4 |
|  | elevated liver function tests | 6 | 4.1 | 3 | 4.2 |
|  | vomiting | 5 | 3.4 | 1 | 1.4 |
|  | miscellaneous gastrointestinal/hepatic event | 24 | 16.3 | 8 | 11.1 |
| Hematologic | any hematologic event | 27 | 18.4 | 12 | 16.7 |
|  | epistaxis | 2 | 1.4 | 2 | 2.8 |
|  | catheter-associated bleeding | 2 | 1.4 | 0 | 0.0 |
|  | anemia | 12 | 8.2 | 7 | 9.7 |
|  | venous thrombosis | 4 | 2.7 | 2 | 2.8 |
|  | arterial thrombosis | 0 | 0.0 | 1 | 1.4 |
|  | suspected pulmonary embolism | 2 | 1.4 | 0 | 0.0 |
|  | confirmed pulmonary embolism | 2 | 1.4 | 0 | 0.0 |
|  | elevated d-dimer | 2 | 1.4 | 0 | 0.0 |
|  | miscellaneous hematologic event | 2 | 1.4 | 0 | 0.0 |
|  | leukocytosis | 8 | 5.4 | 2 | 2.8 |
|  | hemolysis | 1 | 0.7 | 0 | 0.0 |
|  | leukopenia | 1 | 0.7 | 1 | 1.4 |
|  | thrombocytosis | 2 | 1.4 | 0 | 0.0 |
|  | thrombocytopenia | 4 | 2.7 | 1 | 1.4 |
|  | disseminated intravascular coagulation | 1 | 0.7 | 0 | 0.0 |
| Infectious | any infectious event | 29 | 19.7 | 15 | 20.8 |
|  | bacteremia | 3 | 2.0 | 1 | 1.4 |
|  | pneumonia -- bacterial | 16 | 10.9 | 7 | 9.7 |
|  | pneumonia -- unknown | 4 | 2.7 | 4 | 5.6 |
|  | Candida infection without bacteremia | 3 | 2.0 | 1 | 1.4 |
|  | Cytomegalovirus viremia | 0 | 0.0 | 1 | 1.4 |
|  | ventilator-associated pneumonia | 2 | 1.4 | 2 | 2.8 |
|  | urinary tract infection | 3 | 2.0 | 2 | 2.8 |
|  | elevated procalcitonin | 1 | 0.7 | 0 | 0.0 |
|  | indwelling line infection | 1 | 0.7 | 0 | 0.0 |
|  | miscellaneous infectious event | 3 | 2.0 | 1 | 1.4 |
|  | Gastrointestinal infection | 1 | 0.7 | 1 | 1.4 |
| Inflammatory | any inflammatory event | 6 | 4.1 | 3 | 4.2 |
|  | elevated serum/plasma inflammatory marker | 2 | 1.4 | 1 | 1.4 |
|  | fever not attributable to plasma | 4 | 2.7 | 0 | 0.0 |
|  | fever attributable to plasma | 0 | 0.0 | 1 | 1.4 |
|  | hypothermia | 0 | 0.0 | 1 | 1.4 |
| Miscellaneous Adverse Event | any miscellaneous event | 5 | 3.4 | 0 | 0.0 |
|  | fatigue | 1 | 0.7 | 0 | 0.0 |
|  | conjunctivitis | 2 | 1.4 | 0 | 0.0 |
|  | poor appetite | 1 | 0.7 | 0 | 0.0 |
|  | vaginal pain | 1 | 0.7 | 0 | 0.0 |
|  | dizziness | 1 | 0.7 | 0 | 0.0 |
|  | pain (non-specific) | 1 | 0.7 | 0 | 0.0 |
| Musculoskeletal/Dermatologic | any musculoskeletal/dermatologic event | 7 | 4.8 | 5 | 6.9 |
|  | rhabdomyolysis | 2 | 1.4 | 1 | 1.4 |
|  | skin ulcer | 1 | 0.7 | 1 | 1.4 |
|  | urticaria | 1 | 0.7 | 1 | 1.4 |
|  | miscellaneous muscular or dermal | 1 | 0.7 | 2 | 2.8 |
|  | facial swelling unrelated to plasma | 1 | 0.7 | 0 | 0.0 |
|  | hypersensitivity skin rash related to plasma | 1 | 0.7 | 0 | 0.0 |
| Pulmonary | any pulmonary event | 33 | 22.4 | 19 | 26.4 |
|  | acute respiratory failure | 24 | 16.3 | 16 | 22.2 |
|  | hypoxemia without respiratory failure | 1 | 0.7 | 2 | 2.8 |
|  | cough | 1 | 0.7 | 1 | 1.4 |
|  | miscellaneous pulmonary event | 2 | 1.4 | 1 | 1.4 |
|  | pleural effusion | 5 | 3.4 | 2 | 2.8 |
|  | dyspnea | 0 | 0.0 | 1 | 1.4 |
|  | lung atelectasis | 1 | 0.7 | 0 | 0.0 |
|  | pneumothorax | 3 | 2.0 | 0 | 0.0 |
|  | pneumomediastinum | 2 | 1.4 | 0 | 0.0 |
|  | worsening lung opacities on imaging | 2 | 1.4 | 0 | 0.0 |
|  | respiratory tract hemorrhage | 0 | 0.0 | 1 | 1.4 |
|  | bronchospasm | 2 | 1.4 | 0 | 0.0 |
| renal / metabolic | any renal/metabolic event | 32 | 21.8 | 18 | 25.0 |
|  | acute kidney injury | 14 | 9.5 | 7 | 9.7 |
|  | ketoacidosis | 1 | 0.7 | 0 | 0.0 |
|  | metabolic acidosis | 0 | 0.0 | 1 | 1.4 |
|  | hematuria | 1 | 0.7 | 2 | 2.8 |
|  | hyperkalemia | 7 | 4.8 | 0 | 0.0 |
|  | hypernatremia | 6 | 4.1 | 5 | 6.9 |
|  | hypokalemia | 7 | 4.8 | 1 | 1.4 |
|  | hypocalcemia | 1 | 0.7 | 1 | 1.4 |
|  | miscellaneous renal/metabolic event | 2 | 1.4 | 2 | 2.8 |
|  | hyponatremia | 3 | 2.0 | 1 | 1.4 |
| vascular | any vascular event | 1 | 0.7 | 0 | 0.0 |
|  | distal extremity ischemia | 1 | 0.7 | 0 | 0.0 |

**Table S11: Serious adverse events**

| **Serious adverse event** | **Convalescent plasma**  **N=147** | **Control plasma**  **N=72** |
| --- | --- | --- |
| Patients with ≥1 serious adverse event reported - any organ system^a^ | 39 (26.5) | 26 (36.1) |
| Patients with ≥1 serious cardiovascular adverse event | 16 (10.9) | 12 (16.7) |
| Patients with ≥1 serious pulmonary adverse event reported | 24 (16.3) | 16 (22.2) |
| Patients with ≥1 serious renal/metabolic adverse event reported | 7 (4.8) | 5 (6.9) |
| Patients with ≥1 serious infectious adverse event reported | 5 (3.4) | 10 (13.9) |
| Patients with ≥1 serious hematologic adverse event reported | 6 (4.1) | 0 (0.0) |
| Patients with ≥1 serious inflammatory adverse event reported | 1 (0.7) | 0 (0.0) |
| Patients with ≥1 serious gastrointestinal/hepatic adverse event reported | 0 (0.0) | 1 (1.4) |

Legend: ^a^Patients may have had >1 serious adverse event across different organ systems

**Supplemental Figures**

**Figure S1**

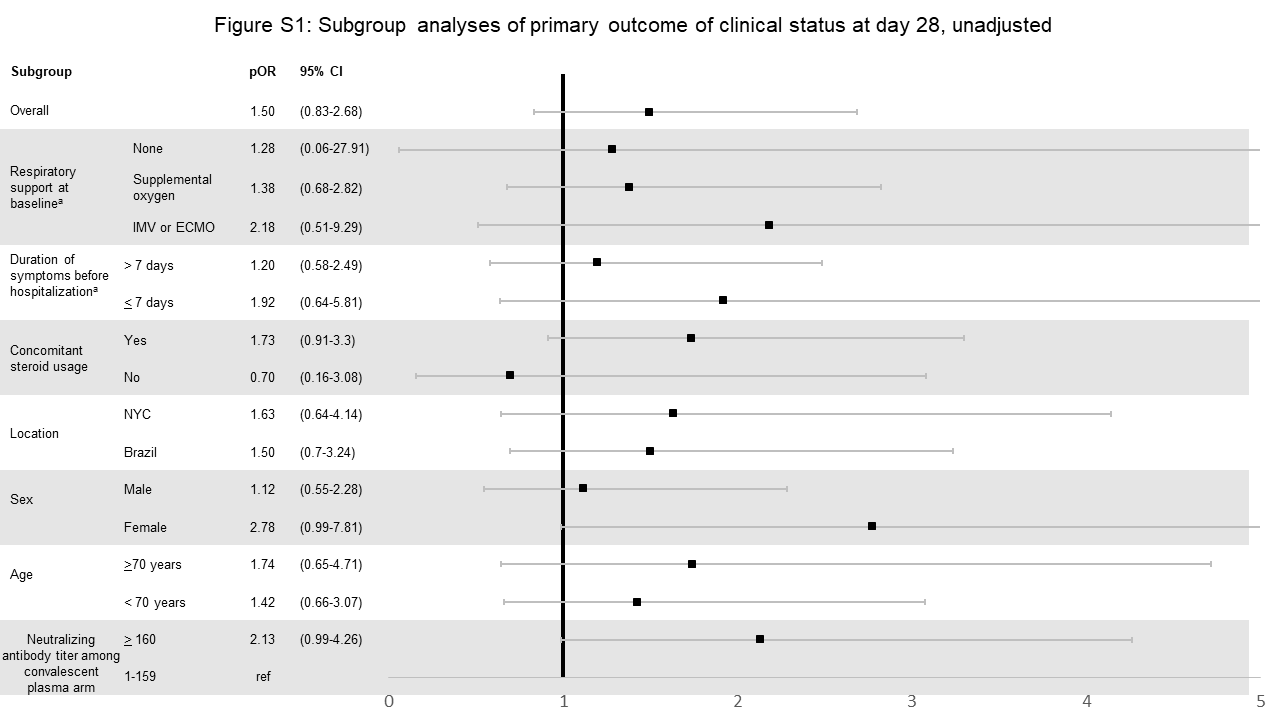

Abbreviations: pOR: proportional odds ratio, IMV: invasive mechanical ventilation, ECMO: extracorporeal membrane oxygenation, NYC: New York City

Legend: ^a^Pre-specified subgroups

**Figure S2**

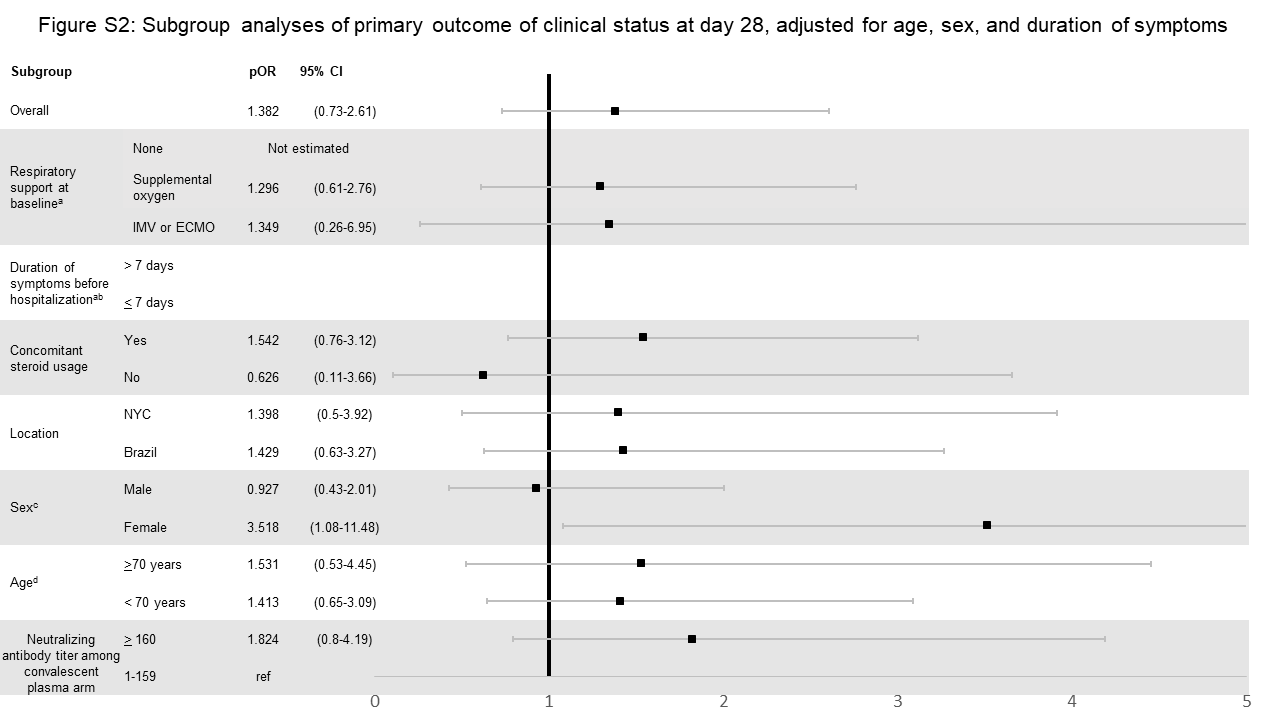

Abbreviations: pOR: proportional odds ratio, IMV: invasive mechanical ventilation, ECMO: extracorporeal membrane oxygenation, NYC: New York City

Legend: ^a^Pre-specified subgroups, ^b^Age- and sex-adjusted estimate presented in Figure 4, ^c^Adjusted for age and duration of symptoms, ^d^Adjusted for sex and duration of symptoms. pOR > 1 associated with improved 28 day clinical status with convalescent plasma.

**Figure S3**

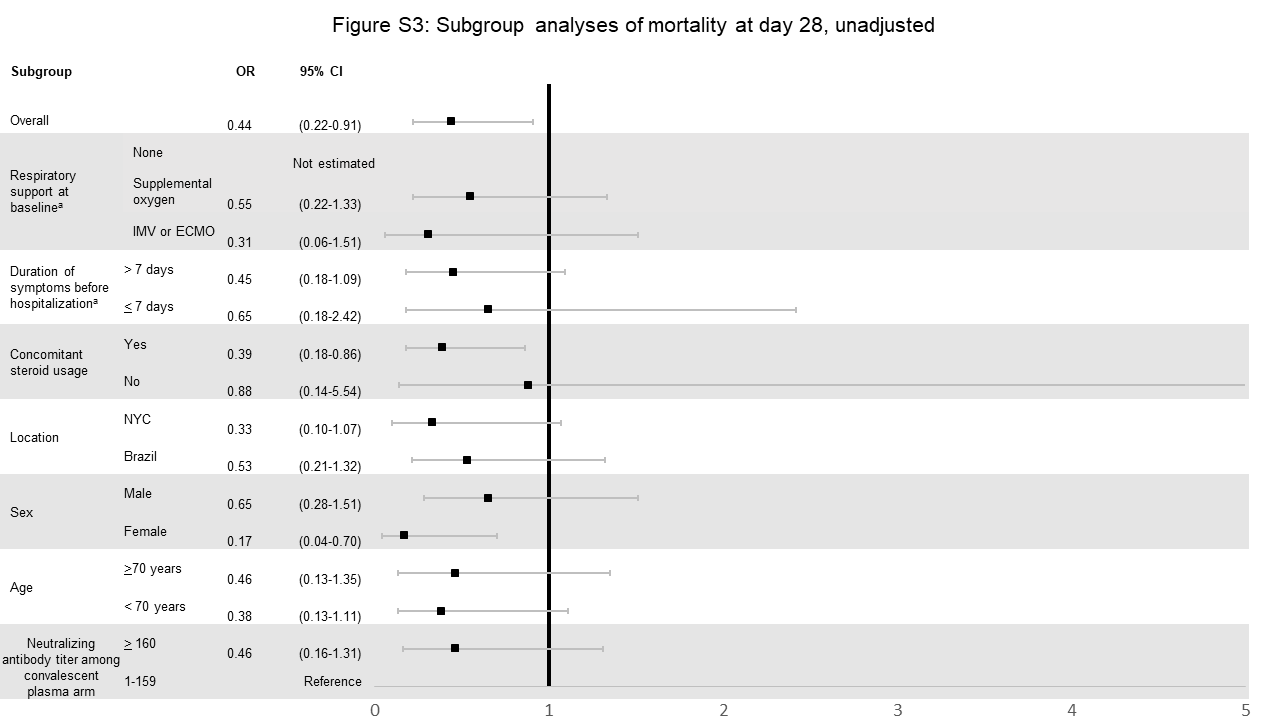

Abbreviations: OR: odds ratio, IMV: invasive mechanical ventilation, ECMO: extracorporeal membrane oxygenation, NYC: New York City

Legend: ^a^Pre-specified subgroups. OR <1 indicates lower 28-day mortality in convalescent plasma group compared to control plasma group.
